# Unraveling Hepatic and Caval Flow to Improve Surgical Planning and Assessment in Fontan Patients via Novel 4D Flow CMR and CFD Methods

**DOI:** 10.1101/2024.12.02.24318116

**Authors:** Akshita Sahni, Vikas Kannojiya, Nicole St. Clair, David M. Hoganson, Peter E. Hammer, Pedro J. del Nido, Rahul H. Rathod, Vijay Govindarajan

## Abstract

The Fontan procedure, a palliative surgery for single-ventricle heart defects, creates a non-physiological circulation that often leads to complications such as pulmonary arteriovenous malformations (PAVMs), thrombosis, and energy loss. As the number of Fontan patients surviving into adulthood continues to grow, precise understanding and improved monitoring of patient-specific hemodynamics is critical for long-term management. Two key challenges hinder progress: 1) the lack of a reliable method to quantify hepatic flow distribution (HFD) from post-operative imaging, a critical determinant of outcomes, and 2) the absence of a validated, patient-specific computational fluid dynamics (CFD) workflow to guide surgical personalization.

This study addresses these gaps by integrating a novel 4D Flow CMR-based particle tracking algorithm with an image-based CFD workflow to quantify and predict Fontan hemodynamics, including HFD, validated with in-vivo data. The 4D Flow-based particle tracking algorithm offers a precise, non-invasive tool for visualizing HFD, demonstrating excellent agreement with phase-contrast MRI (< 5% deviation). Patient-specific CFD models further predicted flow dynamics with high accuracy, validated against in vivo data (< 8% deviation).

Using this integrated workflow, we uncovered uneven mixing between the inferior vena cava and hepatic blood flow, challenging assumptions of uniform mixing and highlighting the critical role of local flow dynamics in determining long-term outcomes. By enabling non-invasive assessment and improved surgical planning, this validated CFD workflow, combined with 4D Flow particle tracking, offers immediate clinical application. With CFD-based surgical planning gaining traction, this approach establishes a new standard for personalized management for patients undergoing treatment for congenital heart diseases.

**Summary:** This study presents a validated workflow integrating 4D Flow CMR and patient-specific CFD to accurately quantify hepatic flow distribution and enhance surgical planning, addressing key challenges in the treatment and long-term management of Fontan patients with single-ventricle disease.

## 1. Introduction

The Fontan operation is a palliative procedure designed for single ventricle heart disease (SHVD), connecting the superior vena cava (SVC), inferior vena cava (IVC), and hepatic veins to the pulmonary arteries via a total cavopulmonary connection using either an extracardiac (EC) conduit or a lateral tunnel (LT) connection (*1*). While lifesaving, this procedure often results in non-physiological blood flow, leading to significant complications such as central venous hypertension, pulmonary arteriovenous malformations (PAVM), cyanosis, ventricular dysfunction, arrhythmia, thrombus formation, stroke, protein-losing enteropathy, exercise intolerance, pleural effusions, and hepatic/renal dysfunction (*2–4*). Efficient blood flow dynamics are critical for the long-term success of the Fontan procedure. Conversely, inefficient fluid dynamics, characterized by disturbed flow, recirculation, and stasis, can cause energy loss and increase the risk of thrombosis, which is particularly problematic in hypercoagulable Fontan patients (*5–7*).

Quantitative visualization of Fontan hemodynamics is essential for clinical teams, enabling them to assess the procedural effectiveness and make informed, data-driven decisions during surgical planning. Accurately visualizing and quantifying total flow distribution (TFD) to the lungs is essential, as any imbalance can lead to inefficient oxygenation and increased cardiac workload, both of which are critical factors affecting the long-term success of the Fontan circulation (*6, 8*). Furthermore, precise assessment of hepatic flow distribution (HFD) is equally important, as imbalanced HFD has been directly linked to the development of pulmonary arteriovenous malformations (PAVMs), a serious complication that can compromise patient outcomes (*9–12*). By evaluating and addressing both total and hepatic flow distribution, clinicians can better predict and mitigate potential complications, ultimately guiding more effective surgical planning. While current imaging modalities such as cardiac catheterization, CT angiography, cardiac magnetic resonance (CMR) imaging, and Doppler ultrasound provide valuable insights, they often lack the ability to quantitatively visualize hepatic flow distribution (HFD) and their mixing with IVC flow due to local fluid dynamics. This limitation hinders the possibility to quantitatively evaluate a Fontan or design an efficient Fontan conduit that ensures balanced HFD with minimized energy loss. Quantitative visualization could significantly improve post-operative evaluation and surgical outcomes.

Patient-specific computational fluid dynamics (CFD) modeling has been previously utilized to provide quantitative insights into Fontan hemodynamics (*6–8, 13, 14*). It is now gaining traction for prospectively design of efficient Fontan pathways that promote optimal flow dynamics (*15, 16*). While these models provide detailed predictions of individual patient hemodynamics (*15–17*), enabling personalized surgical planning, comprehensive validation to ensure their predictive reliability and accuracy has been lacking. Very few studies have attempted to validate patient-specific CFD models, often using in vitro approaches with 3D-printed patient-anatomies and particle image velocimetry (*18*) and MRI data (*19, 20*). While in vitro methods (*18*) provide flow dynamic insights, they are limited to controlled environments, costly and difficult to be integrate into routine clinical practice due to their inability to replicate the complex in vivo conditions. These studies have also made simplifying assumptions, such as using idealized geometries (*18*) or assuming that hepatic flow is fully mixed with IVC flow (*19, 20*) introducing potential inaccuracies. Therefore, there is a critical need for an in-vivo patient-specific validation method and a thoroughly validated CFD model that can accurately replicate the complex flow dynamics within the Fontan circulation, ensuring realistic predictions and enhancing surgical planning outcomes.

Here, we addressed these critical gaps by developing, validating, and applying a novel, cost-effective particle tracking workflow that leverages 4D Flow CMR data to visualize and quantify Fontan hemodynamics with high precision. This workflow enabled the quantitative and visual assessment of hepatic flow distribution (HFD), as well as that of the SVC and IVC flow patterns, establishing its strong clinical relevance. By integrating this CMR-based approach with patient-specific CFD modeling, we achieved comprehensive in-vivo validation of our CFD predictions—marking the first study, to our knowledge, to do so in Fontan patients. We observed highly complex interactions between the SVC flow and the Fontan baffle, contributing to energy loss and uneven pulmonary blood distribution in some patients. Our findings show that IVC and hepatic blood flow do not mix uniformly, as previously assumed, but are instead governed by complex local hemodynamics which challenge conventional understanding of Fontan flow dynamics and underscore the importance of accounting for patient-specific factors in surgical planning. This integrated workflow not only provides an essential tool for evaluating post-operative Fontan circulation but also offers a transformative approach for prospective surgical planning. By delivering critical, patient-specific insights into flow dynamics, this methodology enhances the accuracy of computational workflows, potentially setting a new standard for predictive surgical interventions, ultimately improving clinical outcomes and long-term management for the growing Fontan patient population.

## 2. Results

### 2.1. Validation of 4D Flow CMR-based Particle Tracking Method

To validate our 4D flow particle tracking results (Fig. 1), we quantitatively compared the total flow distribution at the pulmonary arteries with measurements from 2D phase contrast MRI data (Table 1) (*21*). Our analysis showed a remarkable agreement between the flow distribution values obtained from particle tracing and those reported by 2D phase contrast MRI. Specifically, the difference between the two sets of flow distribution values was found to be less than 5% for 4 out of 5 patients in our cohort (Table 1, note that for the fifth patient 2D phase contrast data was not available). This quantitative comparison validates our particle tracking workflow using 4D Flow MRI in the Fontan pathway. Furthermore, the generated flow streamlines, and the particle interaction allowed us to visualize the complex interactions among the SVC, IVC, and hepatic venous flows through the patient’s Fontan pathway to the pulmonary arteries. Tracked particle data delineated flow from individual inlets (IVC, and hepatic veins) and illustrate how they mix in the patient’s Fontan pathway before exiting into the right and left pulmonary arteries (Fig. 1(a)). These findings demonstrate that our 4D flow particle tracking workflow that accurately captured the flow dynamics in Fontan patients, offers a reliable non-invasive method for quantitatively assessing complex caval interactions that can provide post-operative insights into how the surgically designed Fontan pathway diverts individual caval flow in Fontan patients.

**Fig. 1:**
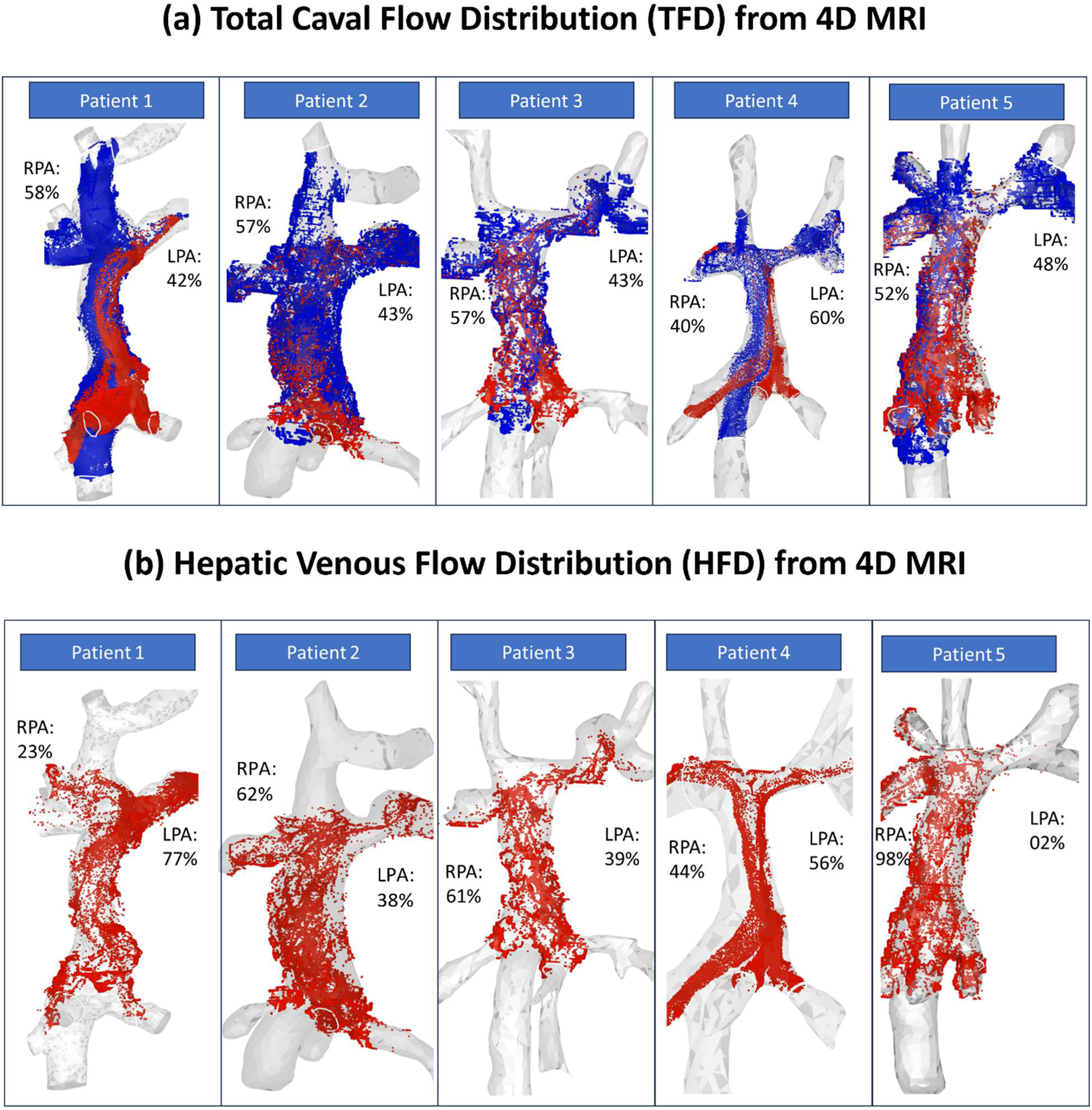
Particle traces illustrating the interactions of flow from each inlet within the Fontan pathway and their distribution to the pulmonary arteries (PAs). Panel (a) shows the total caval flow distribution (TFD) from the superior vena cava (SVC), inferior vena cava (IVC), and hepatic veins, to the right and left pulmonary arteries (RPA, LPA) for patients 1–5 along with the %TFD values. Panel (b) shows the hepatic flow distribution (HFD) to the RPA and LPA for patients 1–5.

**Table 1:** Validation of 4D Flow MRI based particle tracking workflow for total flow distribution (TFD) using 2D phase contrast MRI. . The total flow distribution evaluated using the 4D Flow MRI-based particle tracking technique is shown in the green-highlighted columns, with the difference in TFD compared to the 2D phase contrast MRI results. The findings are consistent with a previous study by Bächler et al.(*45*)

|  |  |  |  | %TFD from 2D<br>phase contrast<br>MRI report |  | %TFD from<br>4D Flow MRI |  | difference in<br>%TFD |
| --- | --- | --- | --- | --- | --- | --- | --- | --- |
| Patient | Age<br>range<br>(yrs.) | n | Total<br>Caval<br>Flow<br>(l/min) | RPA (%) | LPA (%) | RPA (%) | LPA (%) | 2D vs. 4D Flow |
| 1 | 15-20 | 1 | 4.86 | 58 | 42 | 56 | 44 | 2 |
| 2 | 27-32 | 1 | 3.00 | 53 | 47 | 57 | 43 | 4 |
| 3 | 21-26 | 2 | 1.00 | 40 | 60 | 45 | 55 | 5 |
| 4 |  |  | 6.5 | 41 | 59 | 40 | 60 | 1 |
| 5 | 10-15 | 1 | 3.0 | NA | NA | 52 | 48 | - |

### 2.2. Fontan Pathway is Marked by Uneven Flow Distribution and Complex Flow Interactions

Our results revealed highly complex flow patterns at the SVC-Fontan-PA anastomosis that were unique to each patient. Individual contributions of flow from the SVC, IVC, and hepatic veins to the PAs were quantitatively analyzed by our 4D CMR-based particle tracking algorithm. In two out of five patients evaluated (Patients 1 and 2), more than 60% of IVC flow was directed toward LPA while approximately 90% of the SVC flow was directed toward RPA. While Patient 3 had a well-balanced flow distribution, patients 4 and 5 had leftward-skewed SVC and rightward-skewed IVC flow distribution to the PAs (Table 2, Fig. 1(a)). These flow distributions may be attributed to 1) flow collision between SVC and Fontan at the SVC-Fontan-PA anastomosis (*22*); 2) momentum imparted by SVC and Fontan conduit flows that depend on age-based caval flow change (*7, 14*); 3) Fontan pathway curvature, and 4) Fontan-PA anastomosis location. We also noted secondary flow patterns from the SVC-IVC collision in the anastomosis location that can contribute to energy loss in a Fontan (*22*).

**Table 2:**
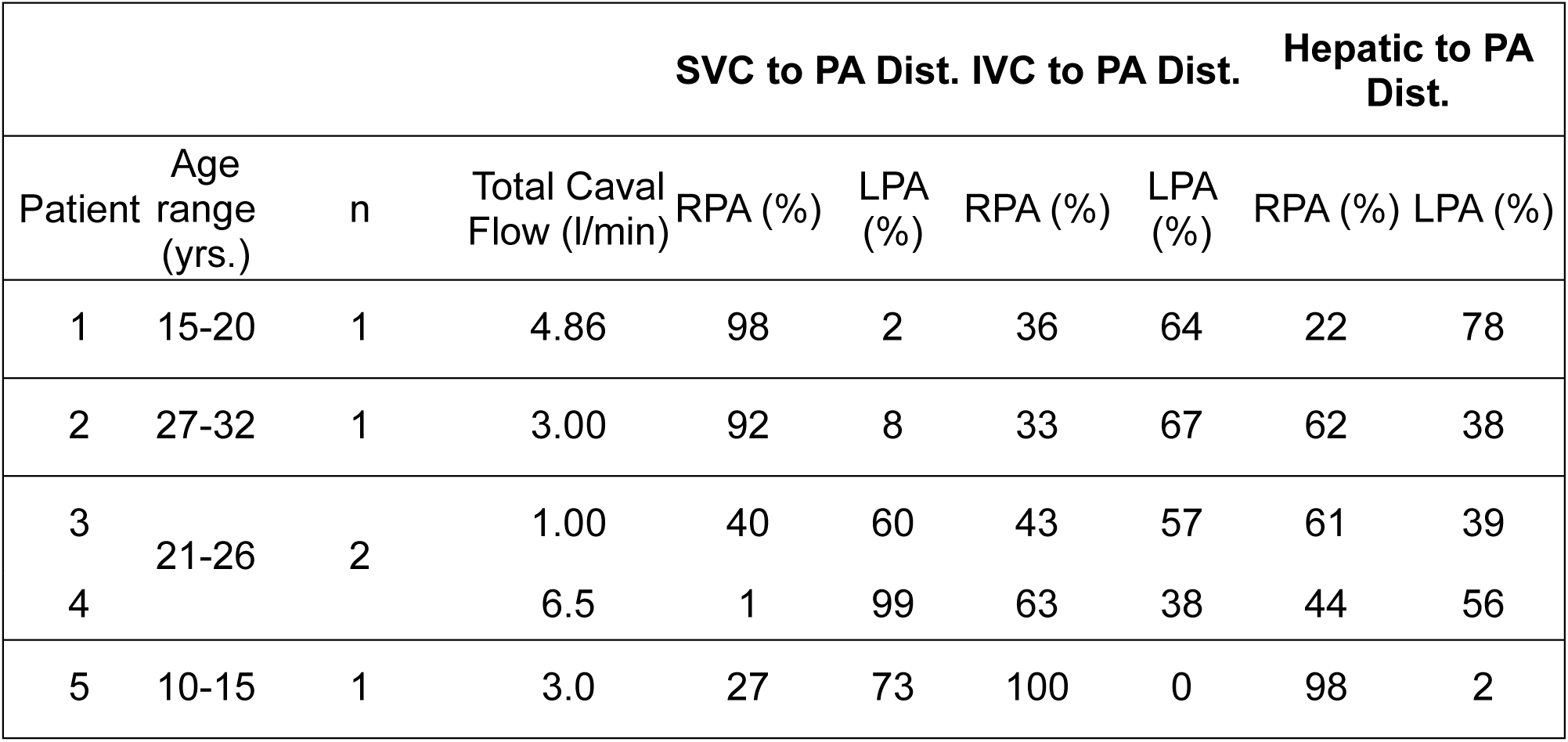
Flow distribution analysis from 4D Flow MRI-based particle tracking, delineating individual contributions from the superior vena cava (SVC), inferior vena cava (IVC), and hepatic veins to the pulmonary arteries (PAs). The variation between IVC flow and hepatic flow distribution is consistent with previous findings by Govindarajan et. al.(*46*) and suggests that hepatic flow is governed by local hemodynamics within the Fontan pathway.

These strong interactions between SVC and Fontan flow as delineated and quantified in this study had a significant impact on hepatic flow distribution in each of the cases. Anatomically, the hepatic veins insert obliquely into the Fontan pathway (Fig. 1). This induced a flow disturbance in the Fontan pathway as hepatic flow interacted with the IVC flow followed by further collision with the SVC flow on its way to the PAs. In all of the patients evaluated, the hepatic flow and IVC flow were not evenly distributed within the Fontan conduit, nor were well mixed as they reached the PAs (Table 2, Fig. 1(b)). This lack of uniform distribution could have critical implications for the development of complications, such as PAVMs.

### 2.3. Predicting Fontan Hemodynamics: Importance of Geometry, Inflow and Outflow

Our patient-specific CFD models were developed after ensuring that the in-flow rates into IVC, hepatics, and SVC, and outflow to the LPA and RPA were consistent with patient’s MRI data. Quantitative as well as qualitative comparisons of local blood flow dynamics under transient flow conditions showed reasonably accurate consistency between model predictions and 4D-Flow CMR data (Fig. 2, Supplementary Figs S1-S8). For all five patient models evaluated, our model predictions of flow evolution using transient flow simulations captured the complex blood flow dynamics within the Fontan pathway as well as the mixing between the IVC blood flow and hepatic blood flow capturing the overall local blood flow dynamics in the Fontan pathway (Fig. 2, Figs S1-S8). Taken together, our results show that local blood flow dynamics within a Fontan baffle that determine a) energy loss and b) hepatic flow distribution (both which determine the efficacy of Fontan via CFD during surgical planning (*16, 23*)) can be accurately predicted by realistic representation of geometry (Fontan conduit, IVC, hepatic veins, SVC and the PAs), inflow and outflow conditions.

**Fig. 2:**
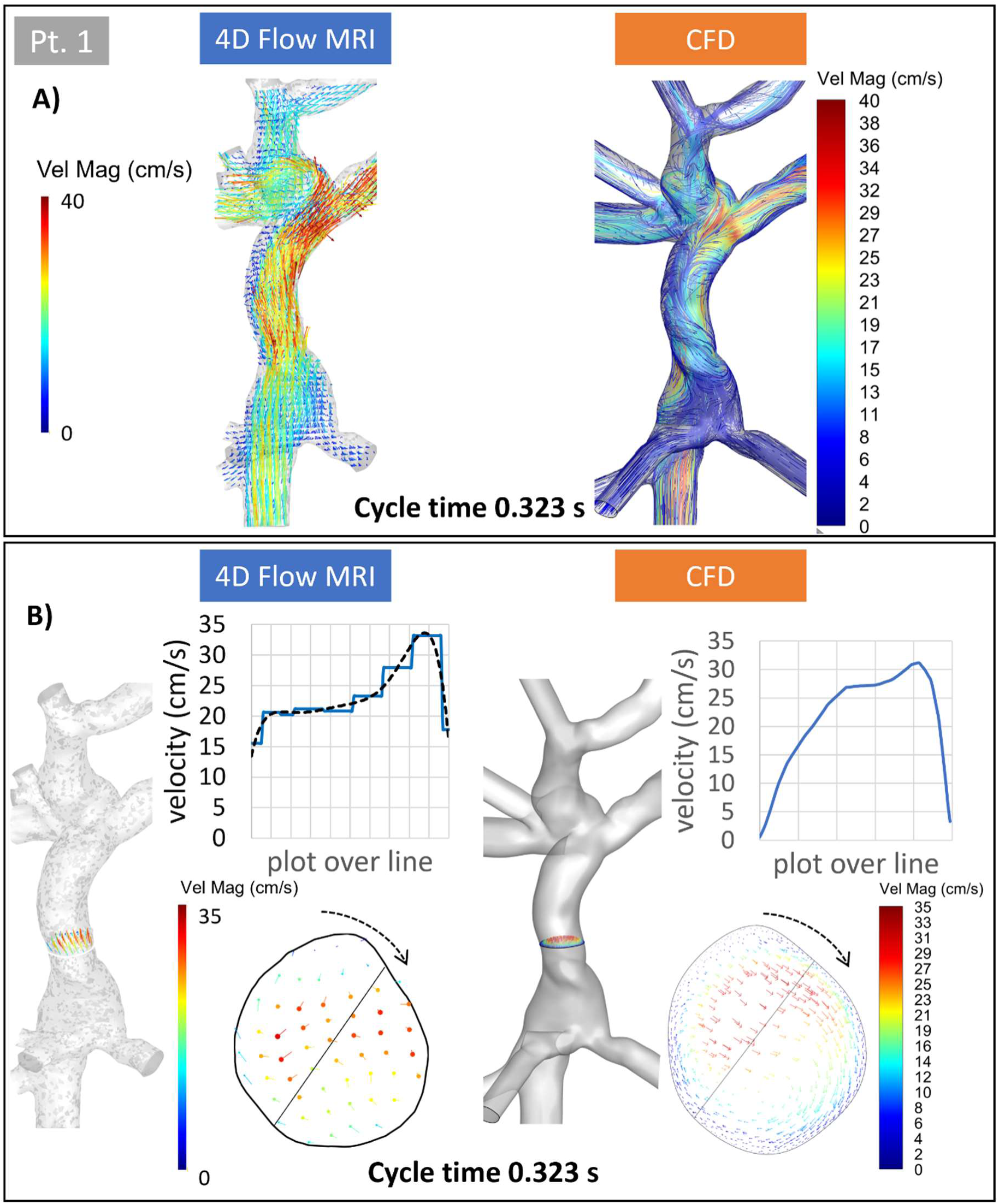
Validation of CFD-predicted local blood flow velocity with 4D Flow MRI for patient 1. Panel A compares Fontan flow velocity streamlines from in-vivo 4D Flow MRI and CFD predictions. Panel B shows similar in-plane flow swirling within the Fontan baffle transverse section (indicated by black arrows) in both methods. Velocity plots along a selected line through the transverse section demonstrate strong agreement between 4D Flow MRI and CFD. *Note: Similar comparisons for patients 2–5 are provided in Supplementary Figures S1–S8*.

Since HFD, quantified using the particle tracking method, is directly measured from the patient’s 4D flow CMR data (Fig 1(b) and Table 2), it serves as an excellent validation source for CFD-predicted hepatic flow distribution in Fontan pathways. We were encouraged to note that CFD-predicted HFD was consistent with the HFD measured by 4D-flow MRI, with a difference of less than 8% (Table 3 and Fig. 3). Qualitatively, the particle trajectories computed from CFD also agreed well with those obtained from the 4D Flow MRI.

**Fig. 3:**
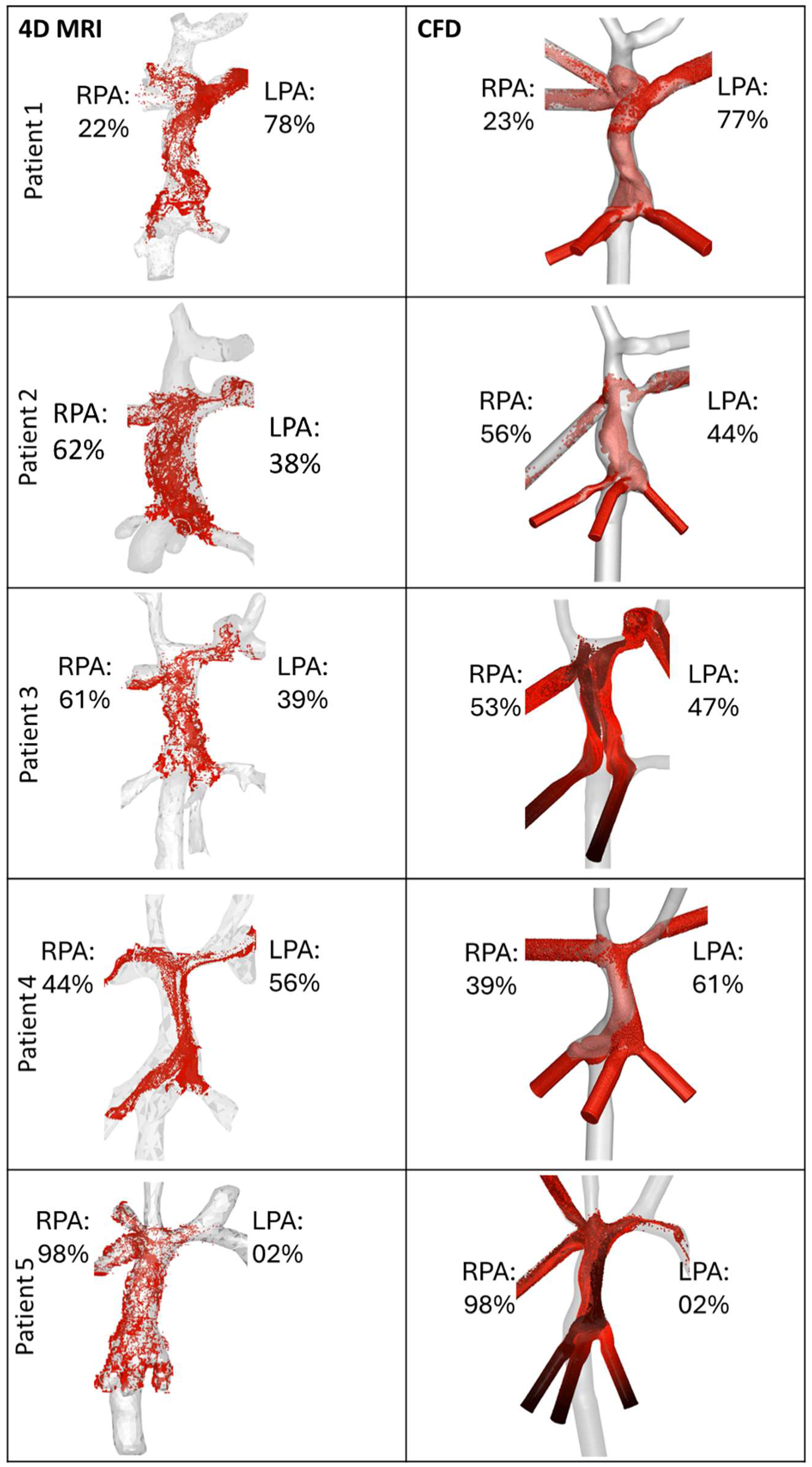
Comparison of hepatic flow distribution between 4D Flow MRI and CFD for each patient at a single snapshot in the cardiac cycle.

**Table 3:**
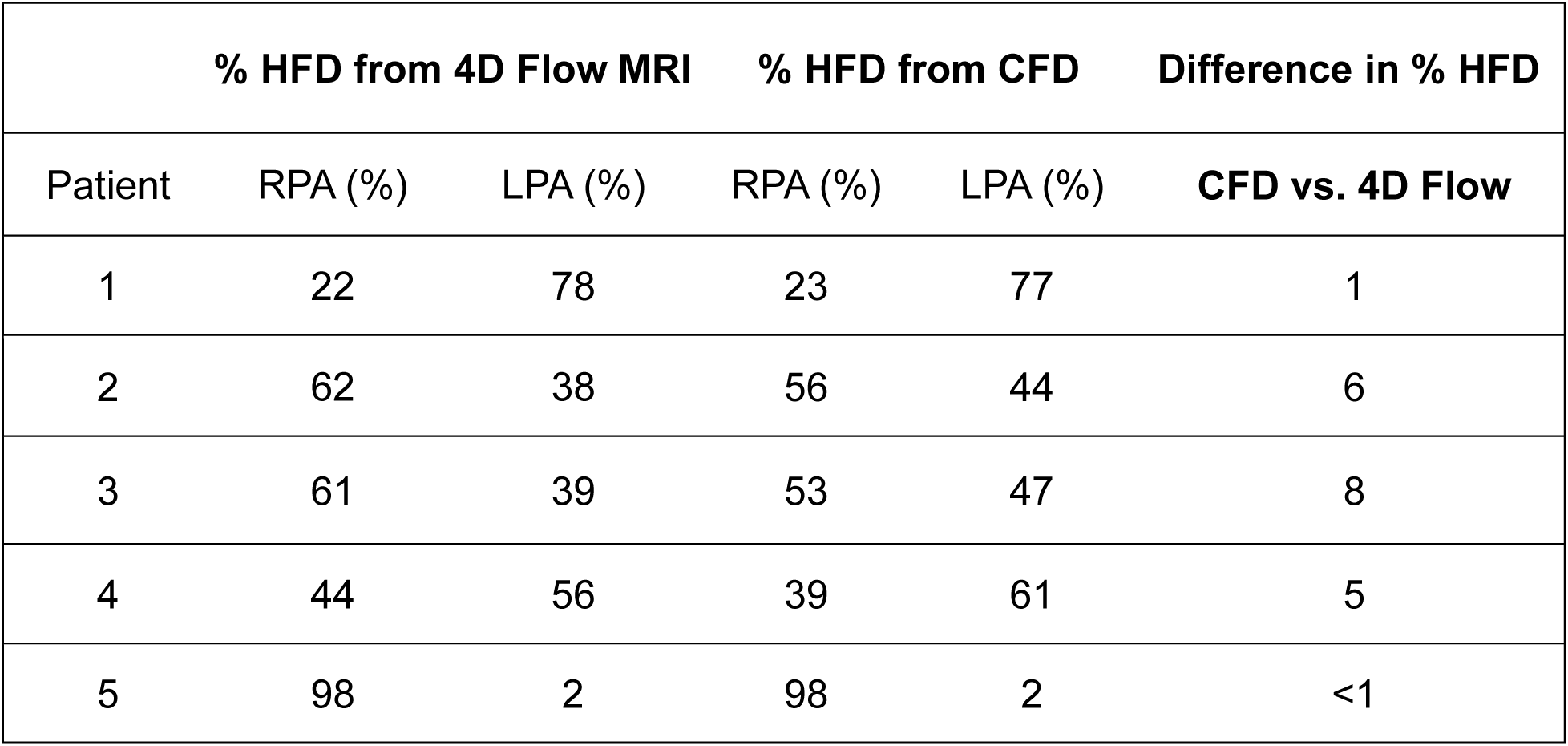
Comparison of CFD-predicted hepatic flow distribution with 4D Flow MRI derived HFD evaluation for patients 1-5.

## 3. Discussion

Efficient blood flow is essential for optimal cardiac function, but it is often negatively impacted by disease, necessitating interventions to restore circulation. In the context of Fontan palliative surgery for patients with single ventricle disease, where an artificial pathway is created for venous blood to reach the lungs, precise assessment of blood flow is vital to predict patient outcomes. This study developed a novel particle tracking-based workflow using 4D flow CMR data to accurately quantify caval flow distributions in Fontan patients, with emphasis on hepatic flow to the lungs, which is crucial for preventing arteriovenous malformations (AVMs) in this patient population. By integrating this method with patient-specific computational fluid dynamics (CFD), we developed a patient-specific approach to validate CFD-based predictions of Fontan hemodynamics with in-vivo data, as demonstrated with our CFD algorithm. Our workflow addressed critical gaps in clinical practice by providing a method for both visualizing and quantifying post-operative caval flow distribution including hepatic flow distribution in Fontan patients. Moreover, to the best of our knowledge, this is the first patient-specific CFD workflow validated comprehensively with in-vivo data, offering a novel approach to improving surgical planning and patient outcomes. Incorporating these validated CFD models into surgical planning not only enhances procedural predictability and refinement but also offers valuable biomechanical insights, potentially setting new standards in the clinical management and long-term outcomes of Fontan patients.

Our results demonstrated a remarkable agreement between the flow distribution values obtained from our 4D flow CMR-based particle tracking workflow and those reported by 2D phase contrast MRI, with differences less than 5% for each patient in our cohort (Table 1). This validated our particle tracking methodology as a reliable non-invasive method for assessing complex venous interactions in the Fontan pathway (Table 1, and Fig. 1). Visualization of individual contributions from the SVC, IVC, and hepatic veins to the pulmonary arteries revealed significant flow patterns and interactions, providing insights into the dynamics at the SVC-Fontan-PA anastomosis (Fig. 1). Our patient-specific CFD model predictions showed a strong correlation with 4D flow CMR data, accurately capturing local blood flow dynamics and hepatic flow distribution for all patients evaluated (Figs 2, 3 and Supplementary Figs. S1-S8), with differences under 8% (Table 3). These findings demonstrate the application of 4D flow CMR-based particle data to establish the validity of patient-specific CFD models, potentially offering a robust, patient-specific tool for optimizing Fontan surgical planning and improving clinical outcomes.

SVC and IVC flow in Fontan patients have several potential clinical implications that impact both the energetics and the hepatic venous return to lungs that play key roles in the success of a Fontan operation. Our previous studies and others have shown that since the time of Fontan completion (age ∼ 3 years), the caval inflow dominance shifts from upper body to lower body as patients reach adulthood (*14, 24*). Using patient-specific CFD studies from a large cohort of extracardiac and lateral tunnel Fontan patients, we demonstrated that this shift can have a profound impact on energy loss and hepatic flow distribution due to a change in local flow dynamics and can be highly patient specific (*7, 14*). Flow disturbances occur at the IVC-Fontan confluence, where IVC and hepatic venous flow transitions from narrow vessels into the expanded Fontan region, leading to reduced velocities, flow separation, and stasis (*7*). This can potentially increase the risk of thrombosis in the hypercoagulable Fontan population (*5, 25, 26*). By accurately visualizing and quantifying the caval flow patterns as shown in our study (Fig. 3), clinicians can gain deeper insights into the hemodynamic efficiency of the Fontan circulation. Moreover, evaluating the flow pattern in a patient’s Fontan can help in identifying regions of disturbed flow, recirculation, and stasis, which are associated with complications such as increased energy loss and thrombus formation.

Hepatic venous return to both lungs is crucial in Fontan circulation, as adequate hepatic blood flow prevents the development of PAVMs (*2, 12, 27*). Clinical studies have shown that PAVMs can resolve following liver transplantation in severe liver disease or redirecting hepatic venous blood to the affected lung in Fontan patients (*9, 28–30*). In Fontan physiology, hepatic blood distribution is influenced by local flow dynamics, the geometry of the pathway, and interactions between caval flows (*10, 19, 22, 31*). Our recent modeling predicted that a shift in caval flow dominance from the SVC to the IVC significantly impacts HFD, with the correlation between IVC flow and HFD decreasing with age (*14*). Rijnberg et al. also predicted that hepatic and IVC flows may not mix well in Fontan (*32*). We were able to confirm these hypothesis using actual patient data with our 4D-flow particle tracking workflow (Table 2). While previous studies have often assumed uniform mixing of IVC and hepatic flow as a single inflow (*6, 17, 33, 34*), our findings (Table S2), along with those of Rijnberg et al. (*32*), indicate that this assumption can lead to significant inaccuracies, highlighting the critical need for patient-specific assessments and a deeper understanding of the complex flow dynamics at the IVC-hepatic confluence.

Given the importance of maintaining balanced HFD in Fontan patients, designing a pathway that achieves this balance and evaluating it quantitatively post-operatively is essential for long-term management of this patient population. In our study, four out of five patients (Patients 1-4) demonstrated a relatively balanced HFD. However, Patient 5 had a highly skewed distribution, with 98% of hepatic flow directed to the RPA and only 2% to the LPA, even though this patient had a balanced total flow distribution of 52% to the RPA and 48% to the LPA (Table 1). Notably, this patient was the only one to exhibit a strong correlation between IVC flow and HFD. Thus, the 4D-flow particle tracking workflow presented here provides a valuable, non-invasive method for post-operative assessment of HFD, offering quantitative insights into hepatic flow dynamics (Table 2). This workflow could potentially be used for identifying and monitoring at-risk patients for any AVM symptoms associated with significantly skewed HFD.

CFD-based surgical planning can provide predictive insights into total flow distribution, HFD, energy loss, pressure gradients, identification of any flow stasis that can contribute to thrombosis, and wall shear stress under multiple flow conditions (*6–8, 14, 35*) and is becoming increasingly popular in designing efficient Fontan pathways (*15, 17, 23*). Our patient-specific workflow incorporated detailed hepatic vein geometry and transient inflow boundary conditions, which more accurately capture the interaction between hepatic and IVC flows as they enter the pulmonary arteries in addition to realistic outflow. These improvements have likely contributed to the strong agreement observed between our CFD model predictions and the 4D-flow-derived HFD (Table 3). These findings suggest that CFD-based surgical planning workflows should incorporate the hepatic vein anatomy and preferably a fully transient flow boundary conditions in order to accurately predict the hemodynamic parameters of interest.

Although this study introduced a novel 4D particle tracking workflow to quantify hepatic and caval flow distributions and used this data to comprehensively validate our patient-specific CFD modeling workflow, some limitations exist. In our 4D Flow MRI-based particle tracing workflow, manually segmenting the Fontan and connected vessels may have introduced variability in the accuracy of the velocity profiles. Since velocity profiles depend on the precise measurement of flow through each vessel, any variation in segmentation could affect the inflow boundary conditions for the CFD simulations. Furthermore, the accuracy of particle tracking was affected by MRI resolution, with motion artifacts sometimes causing stray flow patterns that did not reflect the actual physiology. For our CFD workflow, we assumed rigid walls, based on previous findings from a fluid-structure interaction study that reported minimal effects of conduit wall interactions with blood flow on pressure tracings, hepatic flow distribution, and energy efficiency (*36*). Both quantitative and qualitative comparisons with 4D flow data suggest that this assumption may be valid, at least for extracardiac Fontan constructed with PTFE grafts considered in this study. Future investigations using our 4D flow particle tracking data presented in this study can help determine whether the rigid wall assumption is also valid for lateral tunnel Fontan.

In conclusion, our study demonstrates the efficacy of our novel 4D flow CMR-based particle tracking workflow for quantifying hepatic flow distribution (HFD) and comprehensively evaluating Fontan hemodynamics. The remarkable agreement between our 4D flow CMR-based particle tracking results and 2D phase contrast MRI measurements underscore the reliability of our approach. By integrating this non-invasive imaging technique with patient-specific computational fluid dynamics (CFD) models, we have validated our ability to accurately predict local blood flow dynamics and HFD in Fontan pathways. Quantitative flow data is crucial for early identification of patients at risk for complications, enabling closer monitoring and timely interventions to maintain optimal hemodynamics and prevent late complications. Our findings reveal significant flow patterns and interactions that provide critical insights into the hemodynamic efficiency of the Fontan circulation. This validated methodology offers a robust tool for integrating with existing CFD-based Fontan workflows to optimize hemodynamics, potentially improving clinical outcomes for Fontan patients. Future work will focus on refining this integrated approach into surgical planning workflow to further enhance its clinical utility and predictive accuracy.

## 4. Materials and Methods

### 4.1. Design of 4D-Flow-based Tracking workflow for quantification and visualization of caval flow in Fontan

#### 4.1.1. Vessel Segmentation workflow

The selected patient’s MRI datasets were loaded into GT Flow (GyroTools LLC, Zurich, Switzerland), where the 4D cine series with the maximum number of image slices and phase shifts were chosen to ensure optimal resolution for flow data analysis (Fig. 4A). To segment the vessel lumen, a velocity field mask was created with an image intensity threshold between 100 and 200, effectively suppressing stray velocity vectors in regions without signals. A static 3D mask was then generated using a weighted velocity magnitude map over the vessel volume to define the flow domain. The ‘Region-grow 3D’ feature was employed to seed the lumen with a mask that spans the connected vessel regions. The relevant anatomical structures, including the IVC, Fontan conduit, hepatic veins, SVC, and pulmonary arteries, were manually selected to define the regions of interest (ROI) (Fig. 4B). The segmented 3D vasculature was then smoothened to align with the anatomical blood vessels (Fig. 4B). Subsequently, a surface mesh was created, and centerlines along the vasculature were computed. Circular contours were marked using these centerlines as axes, and vessel segments were identified and named to differentiate each vascular region (Fig. 4B). Finally, an inverse mask of the smoothened vessel segmentation was created to eliminate any artifacts and undesired flow regions.

**Fig. 4:**
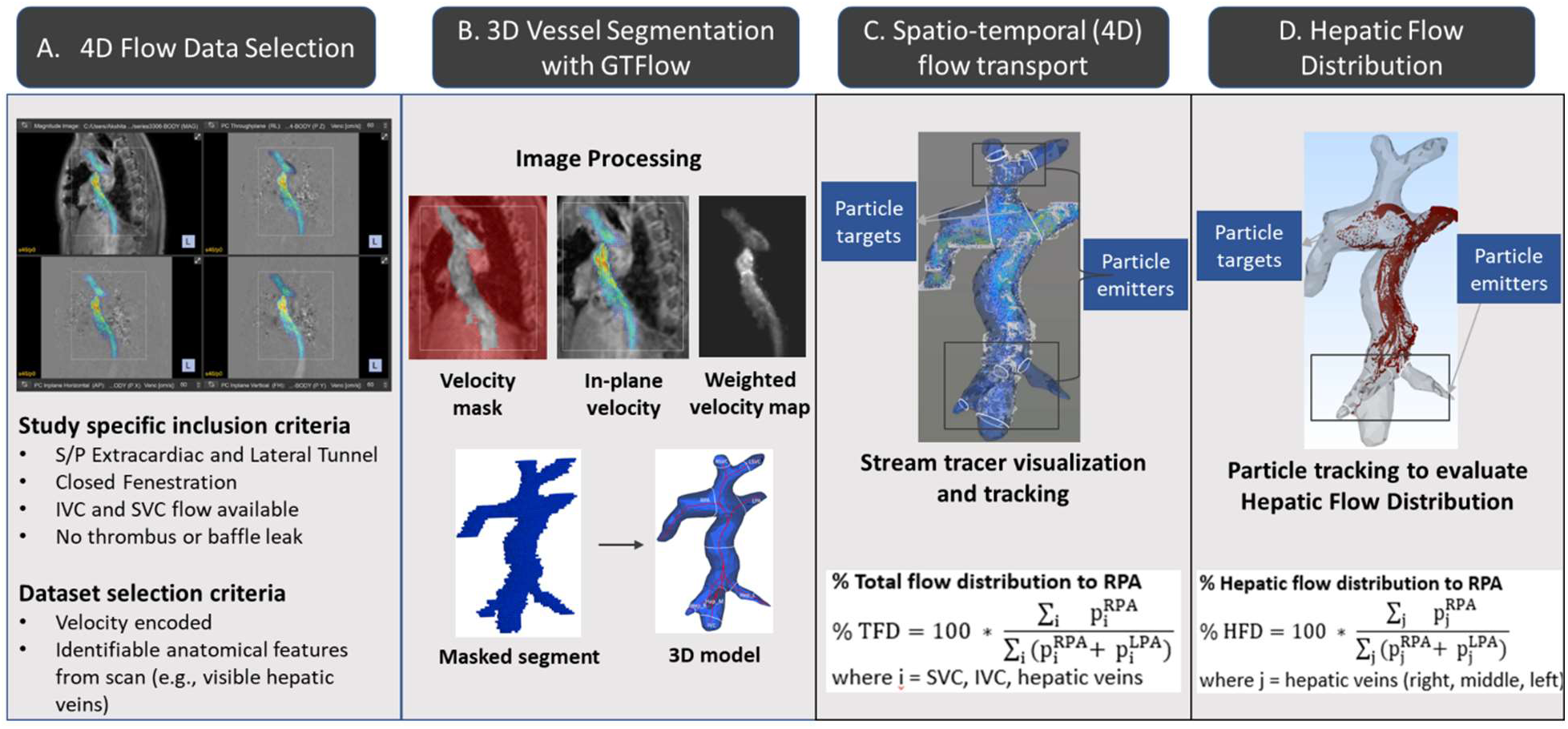
Workflow for 4D Flow MRI data analysis to evaluate hepatic venous flow distribution to the pulmonary arteries (PAs). **A**, 4D Flow MRI data were retrospectively selected from the FORCE study, focusing on a cohort of Fontan patients with closed fenestration and no evidence of Fontan thrombus or baffle leak. **B**, 3D vessel segmentation was performed using GTFlow, reconstructing the Fontan pathway, superior vena cava (SVC), inferior vena cava (IVC), and hepatic veins, which were then converted into a 3D model for quantifying flow distribution. **C**, Particles were released to trace the flow from the SVC, IVC, and hepatic vein inlets into the PAs. The total flow distribution was calculated and validated against 2D phase-contrast MRI data. **D**, After validation of the particle tracking, the hepatic venous flow distribution to the PAs was quantified.

#### 4.1.2. Particle-transport based flow visualization

Massless tracer particles were seeded on each vessel contour (SVC, IVC and hepatic veins) (Fig. 4C). A massless tracer particle transport based on the explicit fourth order Runge-Kutta numerical integration scheme (equation 1) was used to evaluate particle positions with the evolving velocity fields in GT Flow.

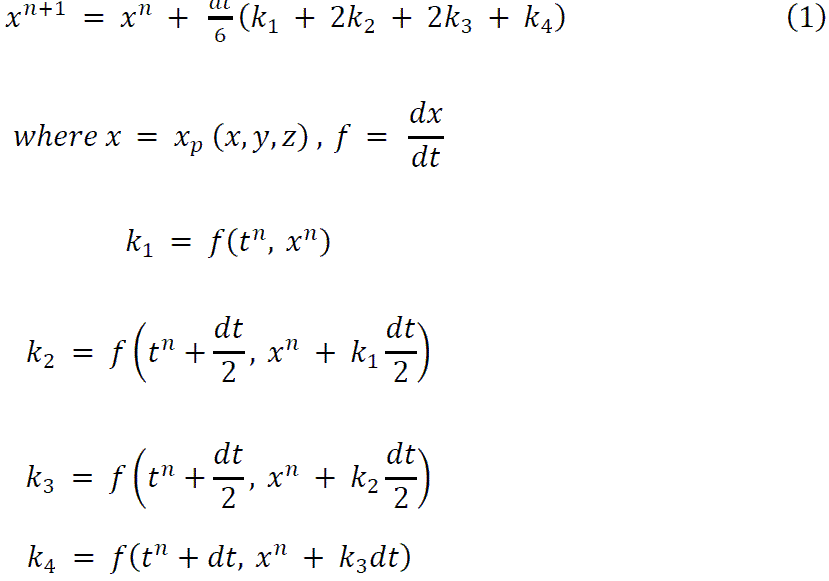

In this 4th-order Runge-Kutta equation *x*^n+l^ is the next position of the particle *p* with *x*_p_ as the current particle position, and it is computed as a weighted sum of four intermediate values (*k*_l_, *k*_2_, *k*_3_, *k*_4_) that approximate the slope at two different time points between (*t*^n^) and *t*^n+l^. Here, each *k*_i_ represents an estimate of the slope at different stages: *k*_l_ is the slope at the current point, *k*_2_ and *k*_3_ are slopes at midpoints using half a time step, and *k*_4_ is the slope at the next full time step.

To optimize the tracer-based flow visualization (Fig. 4D), we chose a seed density factor of 3 and released the seeds 35 times over an interval of 16 ms with a particle trace interval spanning 100 ms to ensure the fate of each particle recorded from the location of its origin to the vessel end. The fluid properties were updated to match those of blood with a density of 1.060 g/cm^3 and a dynamic viscosity of 0.0035 Pa*s. Thus, particles were released from each vessel contour and their trajectory was calculated using an integration time step size 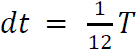, where T is one cardiac cycle.

To compute and visualize the total venous flow to the pulmonary arteries, particles were seeded on Left SVC, Right SVC, IVC, and hepatic veins’ contours and their trajectories were recorded over an interval of 5 cardiac cycles. Similarly, by seeding particles only along the hepatic veins’ contours, the hepatic venous flow to the pulmonary arteries was computed. Throughout the visualization process, vessel opacity was adjusted along with the background color to enhance visibility.

#### 4.1.3. Flow distribution statistics

The SVC, IVC, and hepatic veins were designated as particle emitters, with the PAs serving as target contours for particle tracking. For each emitter-target pair, detailed particle release information, including release time, the number of particles emitted, and the particle counts at each target from each emitter, was obtained from GTFlow. The total caval flow distribution (TFD) to the pulmonary arteries (PAs) was determined by calculating the percentage of particles reaching left pulmonary artery (LPA) and right pulmonary artery (RPA) out of the total particles arriving at both PAs from all inlets. As illustrated in Fig. 1C, particle counts from all emitters (SVC, IVC, and hepatic veins) were obtained at the RPA and LPA targets. The percentage of TFD to the RPA was then computed using the following formula (Equation 2):

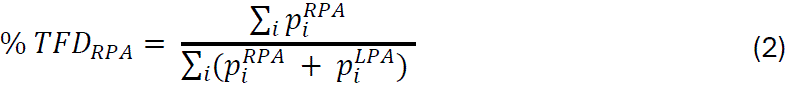

where *i* represents the SVC, IVC, and hepatic veins. The % *T*F*D* _LPA_ is then calculated as 100% − % *T*F*D* _RPA_.

Similarly, to calculate the hepatic flow distribution (HFD) to the RPA, as depicted in Figure 1D, particle counts from hepatic vein emitters (right, middle, and left hepatic) were obtained at the RPA and LPA targets. We then determined the percentage of particles reaching each PA target contour out of the total number of particles reaching both PA target contours from the hepatic vein emitters. The percentage of HFD to the RPA was calculated using a similar formula (Equation 3):

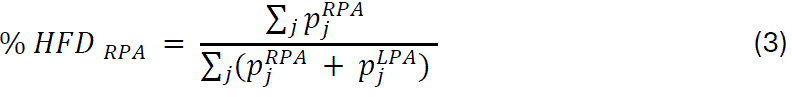

where *j* represents the hepatic veins. The % *H*F*D*_LPA_ is then calculated as 100% −% *H*F*D*_RPA_.

It is important to note that, for the calculation of both total and hepatic venous flow distributions, only particles that reached the PA target contours were considered for the denominators in equations 2 and 3, as not all emitted particles arrived at the target contours. Therefore, comparing the percentage of emitted particles reaching the PAs without this distinction would be misleading.

### 4.2. Design of Computational Flow Models to Predict the Patient’s Fontan Hemodynamics

Our patient-specific workflow for predicting Fontan hemodynamics (Fig. 5) has been developed based on the hypothesis that accurately modeling the physiological anatomy, along with precise inflow and outflow boundary conditions, will enable accurate prediction of local fluid dynamics within Fontan pathways, which determine hemodynamic parameters such as hepatic flow distribution and energy loss. Detailed information on our patient-specific computational flow modeling methodology and its implementation to simulate Fontan hemodynamics is described in our previous studies (*7, 14*) and in the supplementary material.

**Fig. 5:**
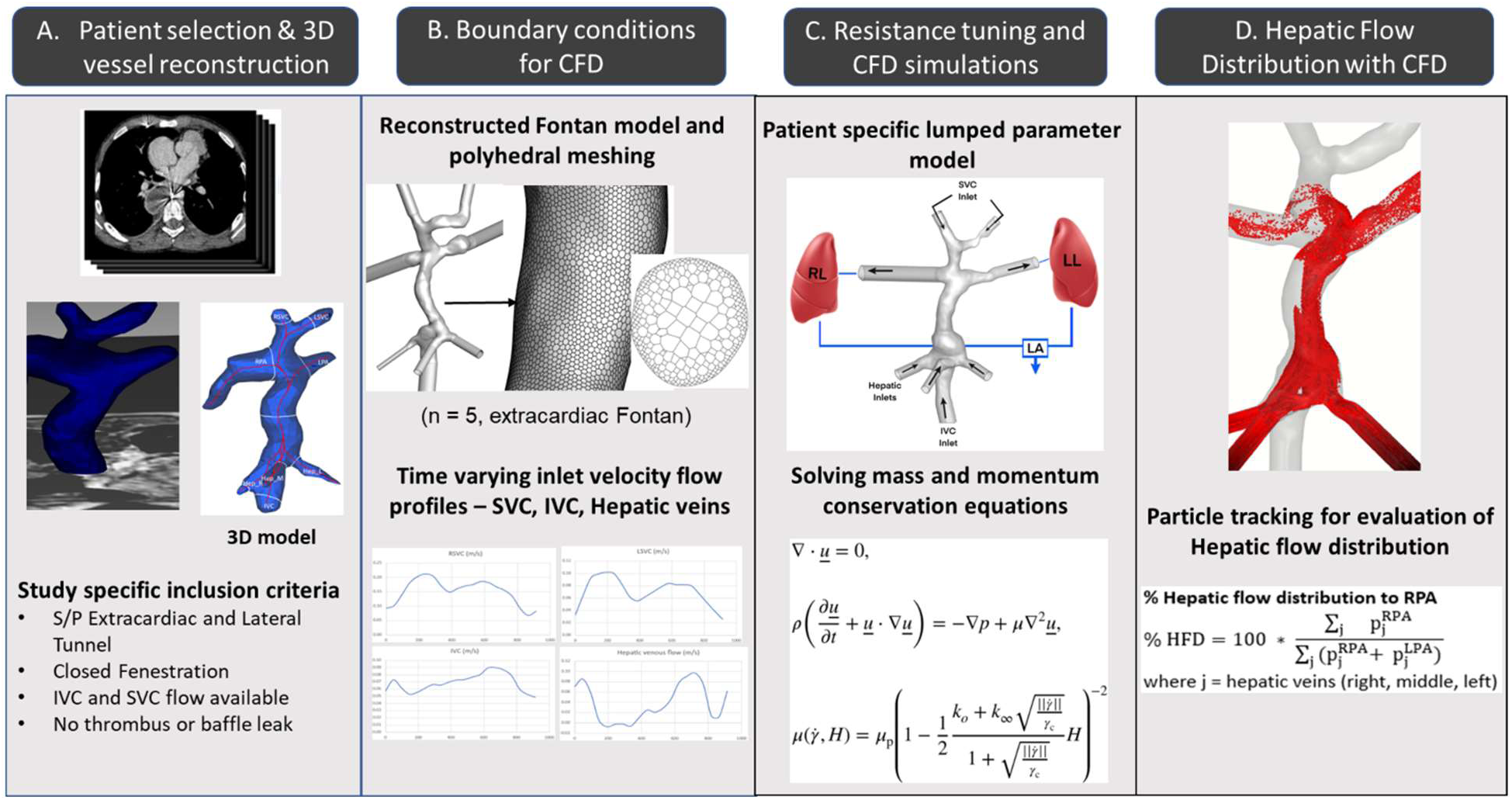
Workflow for CFD analysis to evaluate hepatic venous flow distribution to the pulmonary arteries (PAs). **A**, The patient-specific 3D Fontan anatomy, reconstructed for the 4D Flow analysis workflow, was used for CFD validation, with outlet extensions added to achieve fully developed flow. **B,** Mesh discretization was performed, and fully transient MRI-measured flow rates were applied as boundary conditions. **C**, A lumped parameter model was coupled to the PA outlets, and pulmonary vascular resistance (PVR) was tuned so that the total caval flow distribution to the PAs matched the CMR-measured values within 5%. **D,** Hepatic flow distribution to the PAs was evaluated by seeding massless tracers at the hepatic inlets.

Briefly, our patient workflow incorporated: 1) the 3D reconstructed anatomy of the patient’s Fontan pathway, including the SVC, IVC, LPA, RPA, and hepatic veins, which was segmented from cross-sectional CMR images, smoothed, and adjusted to ensure accurate representation of the patient’s anatomy (*7, 15, 16*); 2) robust octree-based high-density mesh discretization of the computational flow domain performed using ANSYS ICEM(*37*) with proximity-based mesh refinement, ensuring adequate flow resolution to accurately capture boundary layers at the wall (*7, 14, 37*); 3) conversion to a polyhedral mesh (Fig. 5B) known to enhance computational speed by accelerating convergence while maintaining accuracy (*7, 14, 16, 38*); 4) fully transient MRI-measured flow rates applied at the inlets (SVC, IVC, hepatic veins) (Fig. 5B); and 5) a lumped parameter model coupled to the PA outlets (Fig. 2C), which computed outlet pressure based on pulmonary vascular resistance (PVR), that were individually tuned for each patient’s CFD model so that the predicted total flow distribution to the LPA and RPA was consistent with CMR measurements within 5% (*7, 14*). These model features implemented in the CFD software, ANSYS FLUENT(*39*) collectively increased the fidelity of our model predictions. 6) a non-Newtonian Quemada blood viscosity model (*40, 41*) (Fig. 5C) that captures the impact of local shear rate and hematocrit (*7, 14, 40, 41*); previously shown to agree well with in vivo studies (*42*); 7) efficient solution of the incompressible Navier-Stokes equations using the PISO algorithm, which allows for efficient pressure-velocity coupling and stability, and also permits larger time steps for computation; 8) the pressure staggering option (PRESTO!) and a second-order upwind scheme, which ensured accurate pressure gradients and momentum discretization (*7, 14*); The hepatic flow distribution was predicted using massless point particles uniformly seeded at the hepatic vein inlets (Fig. 5D), which were passively carried along with the flow to the PA outlets. These particles were tracked, and their flux quantified as they exited the PA outlets (*10, 14, 33*).

### 4.3. Patient data selection

Four-dimensional phase-contrast magnetic resonance imaging (4D Flow MRI) data for Fontan patients was retrospectively collected from our center and the FORCE database (*43, 44*). This study utilized anonymized and de-identified CMR data obtained as part of routine clinical care, with approval from the Institutional Review Board at Boston Children’s Hospital (Protocol title: Utilizing Patient-Specific Computational Fluid Dynamics Modeling to Optimize the Fontan Procedure), with a waiver of informed consent. Patients were selected based on the following criteria: absence of thrombosis in the Fontan pathway, pulmonary arteriovenous malformations (PAVMs), interrupted IVC with azygous continuation, baffle leak, open fenestration, and aortopulmonary collateral flow greater than 40%. The patient datasets were also checked to ensure velocity encoding in three directions, enabling comprehensive flow analysis within the 3D vasculature (Fig. 5A). Additionally, it was confirmed that the CMR data had sufficient image resolution and clear delineation of hepatic venous flow.

## Disclosure

### Author Contributions

AS: Development and implementation of 4-D particle tracking workflow, CFD simulations, manuscript methodology, and analysis of results; VK: CFD simulations, analysis of CFD results; NS: Data curation; VG: Conceptualization, study design, analysis, and development of CFD workflow, and manuscript draft; All authors analyzed the results, reviewed, and edited the manuscript.

## Data Availability

All data produced in the present study are available upon reasonable request to the authors

## Acknowledgements

Research reported in this publication was supported by the National Heart, Lung, And Blood Institute of the National Institutes of Health under Award Number R01HL161507. The content is solely the responsibility of the authors and does not necessarily represent the official views of the National Institutes of Health. Authors also acknowledge the computational resources provided by the Texas Advanced Computer Center for performing the flow simulations as a part of this study. Authors also acknowledge Lauren Marshall MS for segmentation of a patient-specific Fontan anatomy.

## Ethics Statement

This study was approved by the Institutional Review Board at Boston Children’s Hospital (Protocol Title: Utilizing Patient-Specific Computational Fluid Dynamics Modeling to Optimize the Fontan Procedure). The IRB waived the requirement for informed consent as all data were anonymized and collected as part of routine clinical care.

## Supplementary Materials

Supplementary Figures S1-S8

## Supplementary Figures

**Fig. S1:**
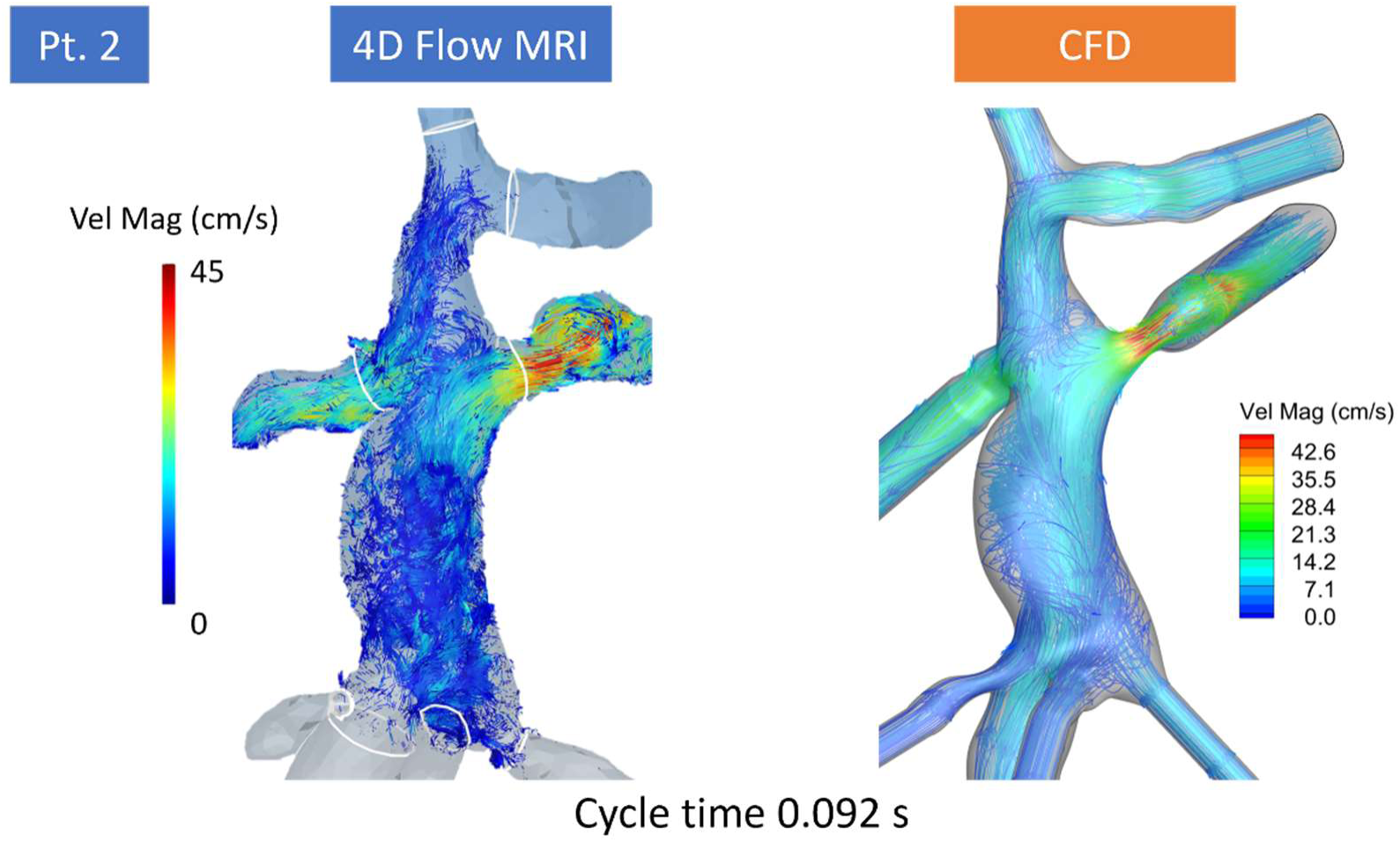
Validation of CFD-predicted local flow velocity with 4D Flow MRI for patient 2. Comparison of Fontan flow velocity streamlines over the volumetric flow domain from in-vivo 4D Flow MRI and CFD-predicted flow.

**Fig. S2:**
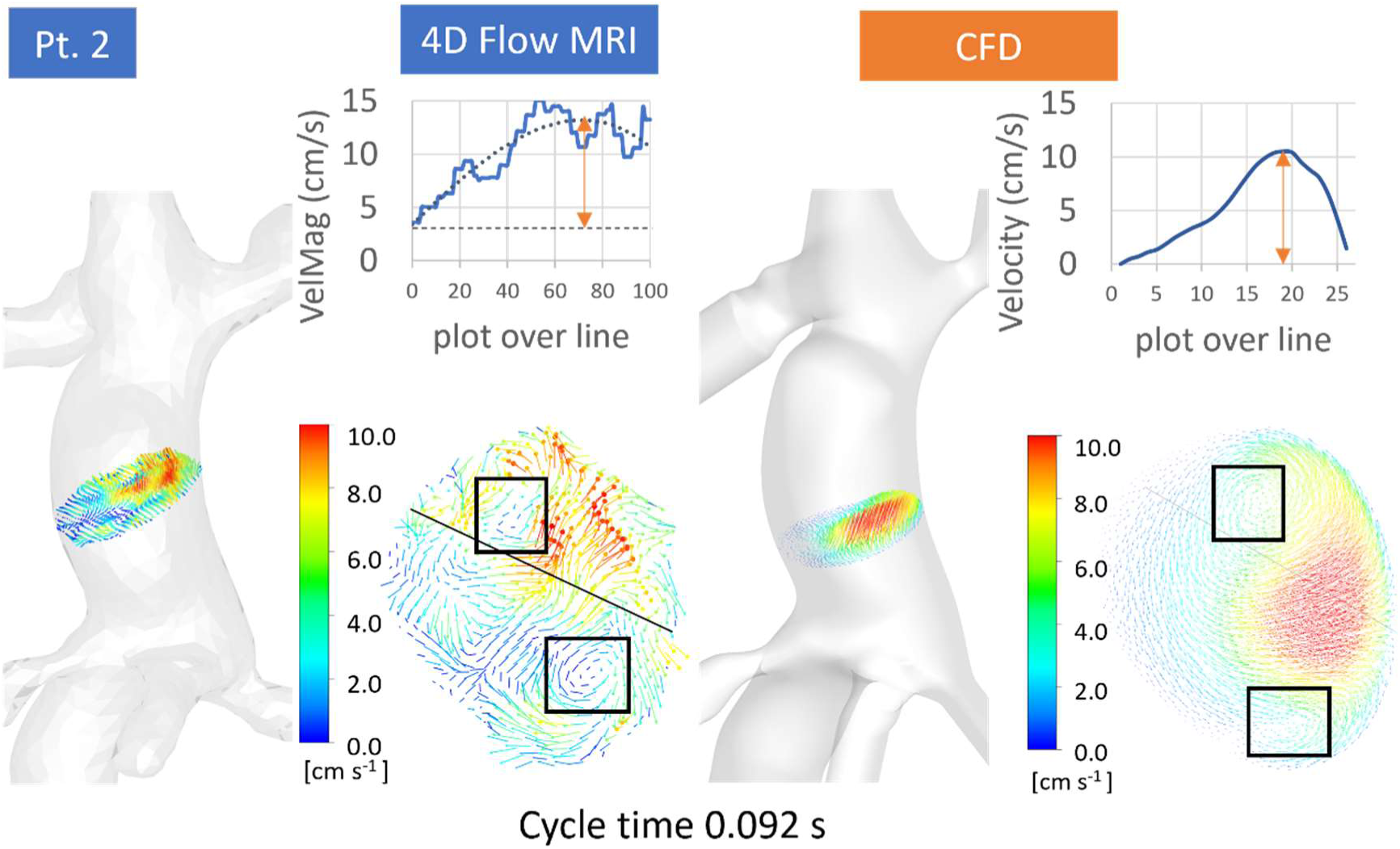
Validation of CFD-predicted local flow velocity with 4D Flow MRI for patient 2. Note the similarity between secondary vortices arising within the Fontan baffle transverse section (indicated by black boxes) in both methods. Velocity plots along a selected line through the transverse section also demonstrate strong agreement between 4D Flow MRI and CFD.

**Fig. S3:**
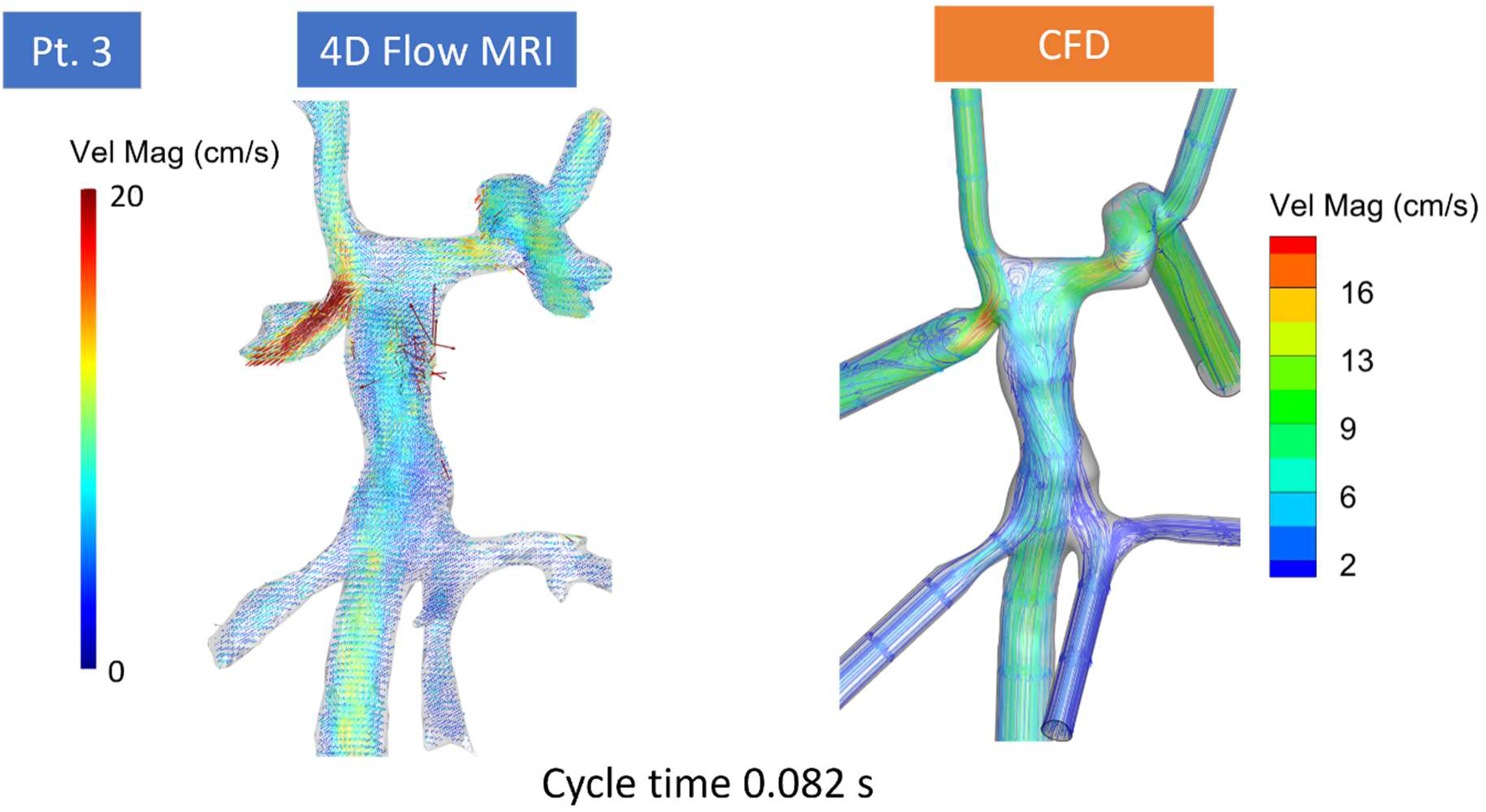
Validation of CFO-predicted local flow velocity with 40 Flow MRI for patient 3. Comparison of Fontan flow velocity streamlines over the volumetric flow domain from in-vivo 4D Flow MRI and CFO-predicted flow.

**Fig. S4:**
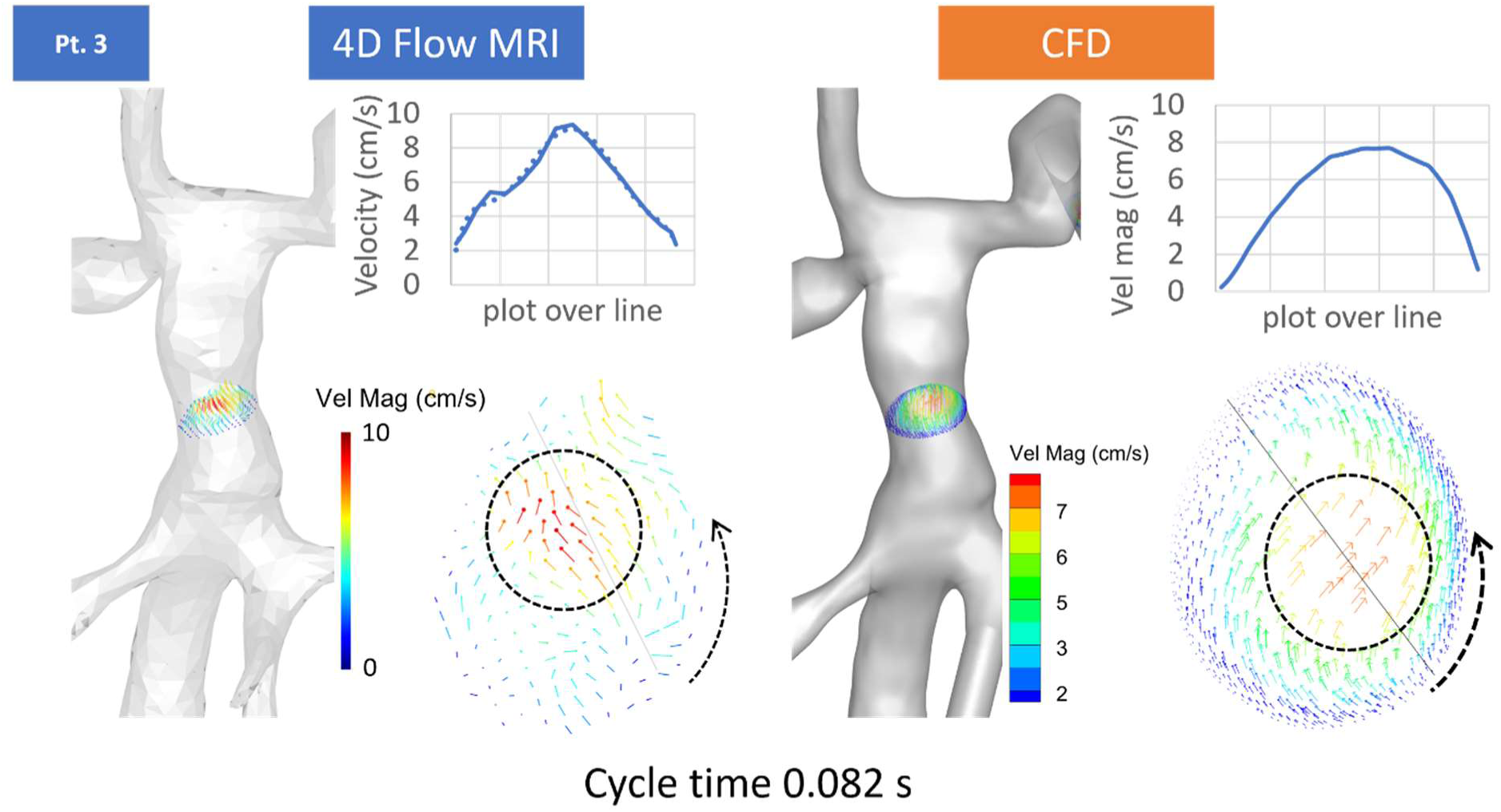
Validation of CFO-predicted local flow velocity with 40 Flow MRI for patient 3. Note the similarity between the high velocity regions arising within the Fontan baffle transverse section (indicated by black circles) in both methods. Velocity plots along a selected line through the transverse section also demonstrate strong agreement between 4D Flow MRI and CFD.

**Fig. S5:**
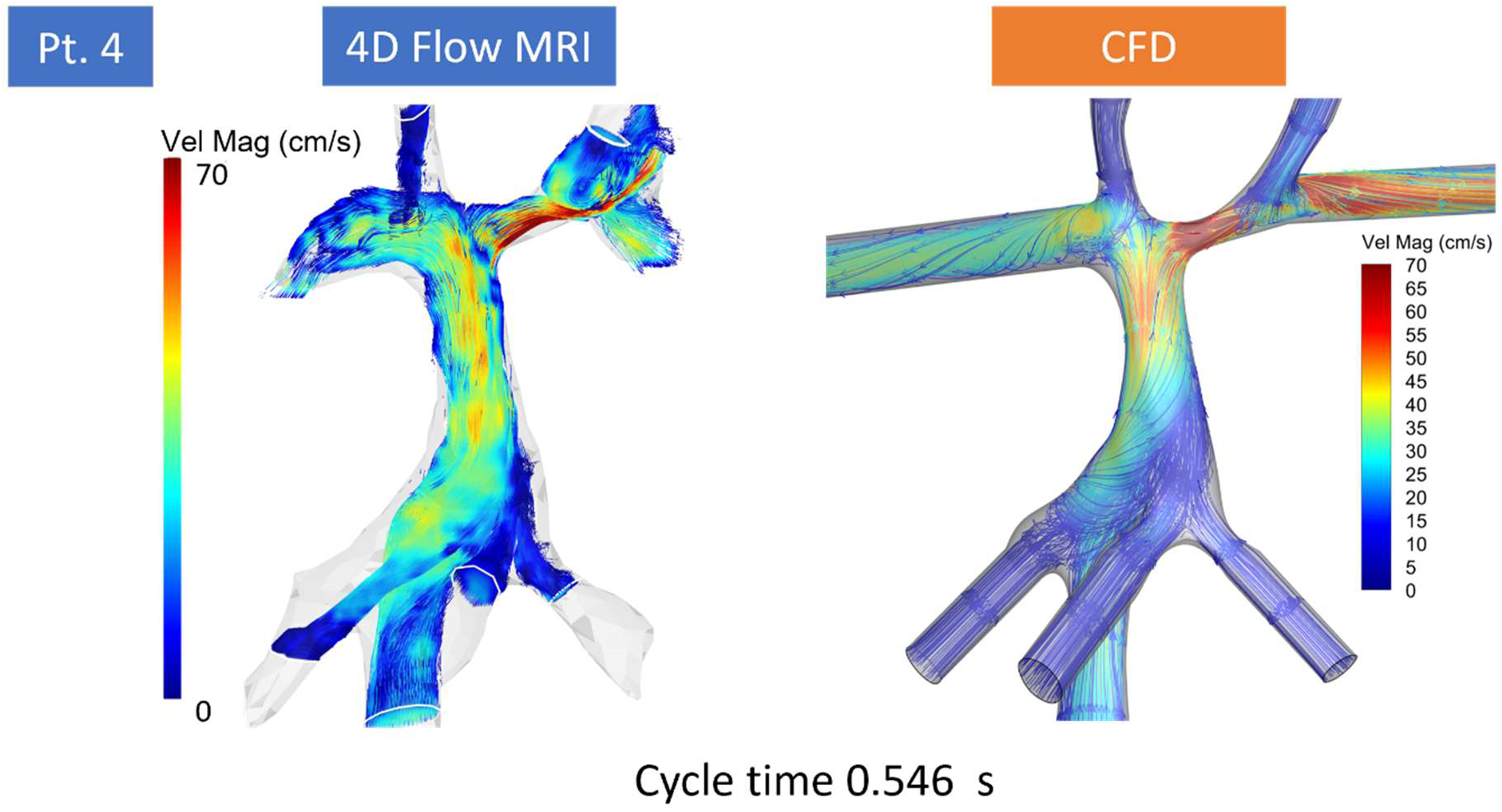
Validation of CFO-predicted local flow velocity with 40 Flow MRI for patient 4. Comparison of Fontan flow velocity streamlines over the volumetric flow domain from in-vivo 4D Flow MRI and CFO-predicted flow.

**Fig. S6:**
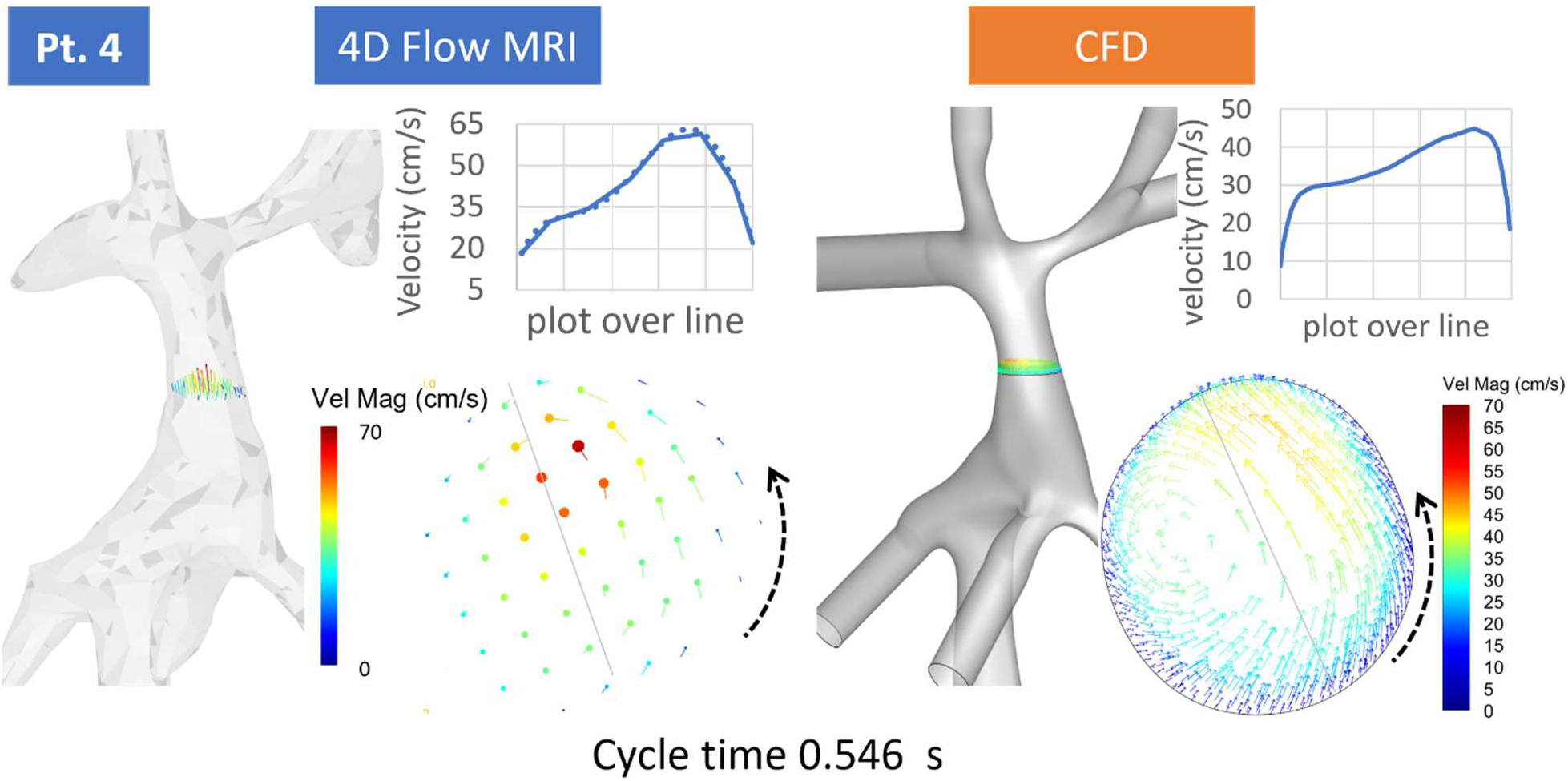
Validation of CFO-predicted local flow velocity with 40 Flow MRI for patient 4. Note the similarity between the in-plane flow swirling arising within the Fontan baffle transverse section (indicated by black arrows) in both methods. Velocity plots along a selected line through the transverse section also demonstrate strong agreement between 4D Flow MRI and CFD.

**Fig. S7:**
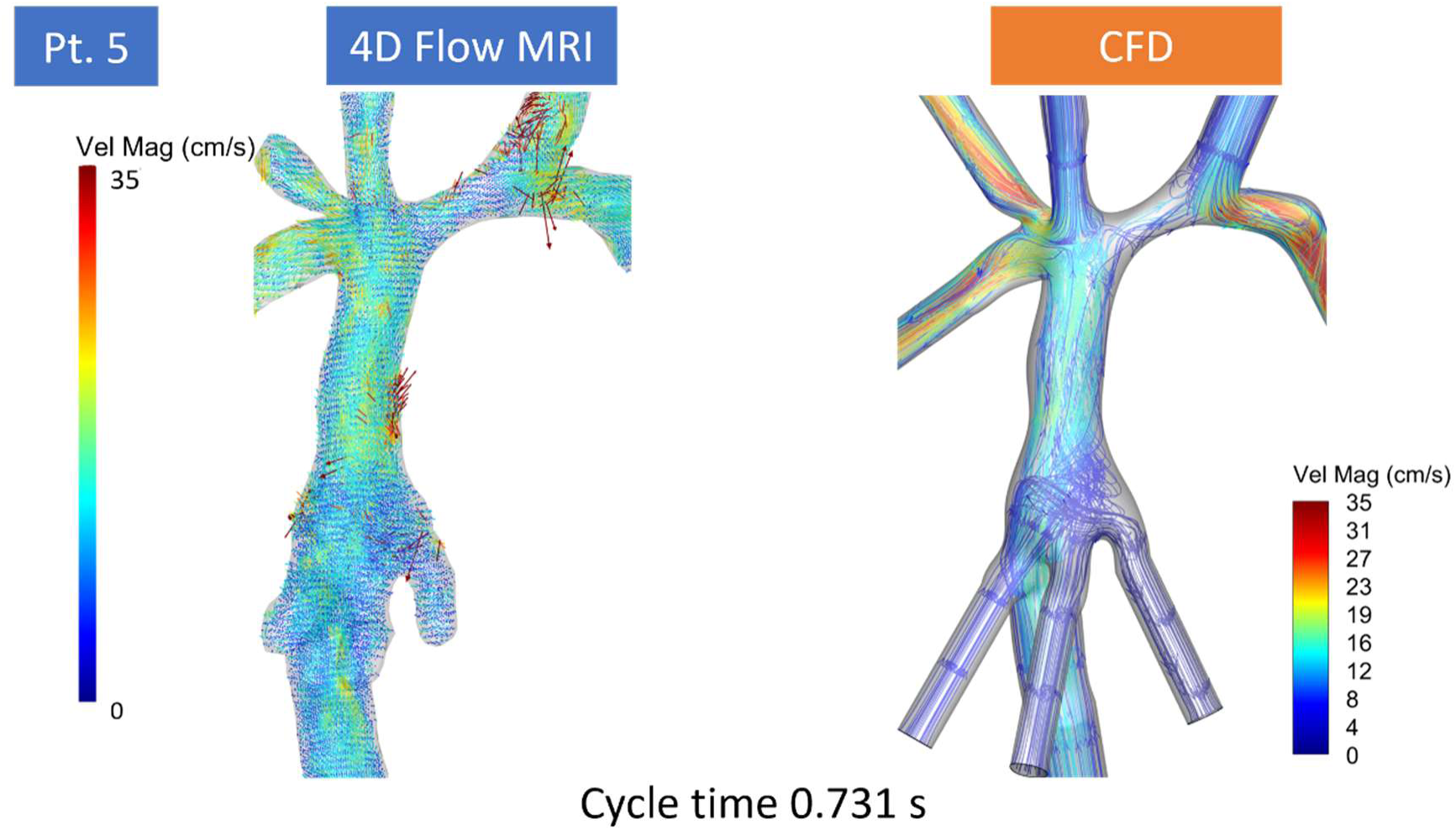
Validation of CFO-predicted local flow velocity with 40 Flow MRI for patient 5. Comparison of Fontan flow velocity streamlines over the volumetric flow domain from in-vivo 4D Flow MRI and CFD-predicted flow.

**Fig. S8:**
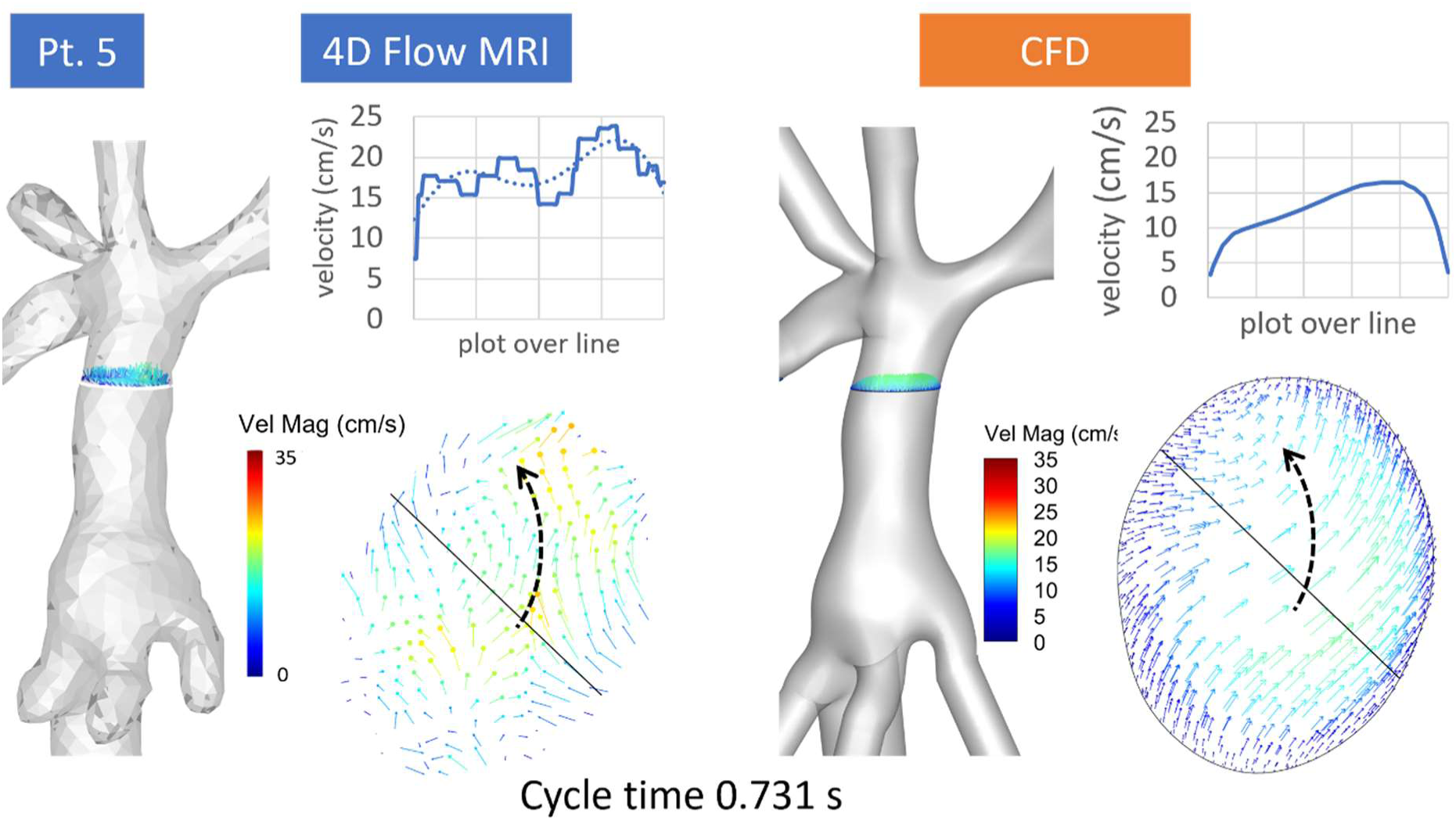
Validation of CFO-predicted local flow velocity with 40 Flow MRI for patient 5. Note the similarity between the in-plane flow swirling arising within the Fontan baffle transverse section (indicated by black arrows) in both methods. Velocity plots along a selected line through the transverse section also demonstrate strong agreement between 4D Flow MRI and CFD.

## FIG

